# Investigation of subsequent and co-infections associated with SARS-CoV-2 (COVID-19) in hospitalized patients

**DOI:** 10.1101/2020.05.29.20117176

**Authors:** Matthew P. Crotty, Ronda Akins, An Nguyen, Rania Slika, Kristen Rahmanzadeh, Marie H. Wilson, Edward A. Dominguez

**Author notes:** **Correspondence to:** Matthew Crotty, Department of Pharmacy, Methodist Dallas Medical Center, Dallas, Texas, USA.

## Abstract

**Background:** SARS-CoV-2 has drastically affected healthcare globally and causes COVID-19, a disease that is associated with substantial morbidity and mortality. We aim to describe rates and pathogens involved in co-infection or subsequent infections and their impact on clinical outcomes among hospitalized patients with COVID-19.

**Methods:** Incidence of and pathogens associated with co-infections, or subsequent infections, were analyzed in a multicenter observational cohort. Clinical outcomes were compared between patients with a bacterial respiratory co-infection (BRC) and those without. A multivariable Cox regression analysis was performed evaluating survival.

**Results:** A total of 289 patients were included, 48 (16.6%) had any co-infection and 25 (8.7%) had a BRC. No significant differences in comorbidities were observed between patients with co-infection and those without. Compared to those without, patients with a BRC had significantly higher white blood cell counts, lactate dehydrogenase, C-reactive protein, procalcitonin and interleukin-6 levels. ICU admission (84.0 vs 31.8%), mechanical ventilation (72.0 vs 23.9%) and in-hospital mortality (45.0 vs 9.8%) were more common in patients with BRC compared to those without a co-infection. In Cox proportional hazards regression, following adjustment for age, ICU admission, mechanical ventilation, corticosteroid administration, and pre-existing comorbidities, patients with BRC had an increased risk for in-hospital mortality (adjusted HR, 3.37; 95% CI, 1.39 to 8.16; P = 0.007). Subsequent infections were uncommon, with 21 infections occurring in 16 (5.5%) patients.

**Conclusions:** Co-infections are uncommon among hospitalized patients with COVID-19, however, when BRC occurs it is associated with worse clinical outcomes including higher mortality.

## Background

SARS-CoV-2 is a pandemic coronavirus that has drastically affected healthcare globally and causes COVID-19, a disease that is associated with substantial morbidity and mortality. [1–4] COVID-19 is predominantly a respiratory disease and frequently causes pneumonia. [1–4]

Co-infections have been described between other respiratory viruses and bacterial pathogens, sometimes leading to worse clinical outcomes. [5, 6] Early reports suggest that co-infections between SARS-CoV-2 and other respiratory pathogens (both viral and bacterial) occur at varying rates and with an unknown impact on clinical outcomes. [7] In contrast, empiric antibiotic use has been reported among most patients, including broad spectrum agents and those with potential toxicities. [1, 8, 9] Moreover, patients with COVID-19 frequently have prolonged hospitalizations (including intensive care unit [ICU] stays) and often require mechanical ventilation and other invasive procedures that put them at a high risk for nosocomial infections. [1–3]

Among currently available descriptions of co-infections in patients with COVID-19, most are lacking details related to diagnostic work-up, critical illness, time to infection detection, pathogens identified, and clinical outcomes. [7] A more detailed description and evaluation of co-processes may allow for a better understanding of the disease process and patient prognosis, as well as inform improved antimicrobial stewardship practices. We sought to describe rates and pathogens involved in co-infection or subsequent infections and their impact on clinical outcomes among hospitalized patients with COVID-19.

## Methods

### Study Population and Data Collection

The Aspire institutional review board approved this multicenter observational cohort study as minimal-risk research using data collected for routine clinical practice and waived the requirement for informed consent. Patients age 18 years or older admitted to any of the 4 acute care hospitals within Methodist Health System in Dallas, Texas, USA between March 1, 2020, and April 30, 2020 were eligible for inclusion. All consecutive patients who were sufficiently medically ill to require hospital admission with confirmed SARS-CoV-2 infection by positive result on reverse-transcriptase-polymerase-chain-reaction (RT-PCR) testing of a nasopharyngeal during the index admission or in the emergency department prior to admission were included. Patients were excluded if either they only received care in the emergency department or they were admitted to inpatient/observation status for less than 24 hours. Clinical outcomes were monitored until May 14, 2020, the final date of follow-up. All data were collected from the electronic health record (Epic; Verona, Wisconsin; http://www.epic.com).

Data collected included patient demographic information, comorbidities, initial laboratory tests, diagnoses during the hospital course, inpatient medications, treatments (including invasive mechanical ventilation), and outcomes (including length of stay, discharge, and mortality). Demographics, baseline comorbidities, and presenting clinical studies were available for all admitted patients. All clinical outcomes are presented for patients who completed their hospital course at study end (discharged alive or dead). Clinical outcomes available for those in hospital at the study end point are presented, including invasive mechanical ventilation, ICU care, and length of stay in hospital. Outcomes such as discharge disposition and readmission were not available for patients in hospital at study end because they had not completed their hospital course. Initial laboratory testing was defined as the first test results available, typically within 24 hours of admission. For initial laboratory testing and clinical studies for which not all patients had values, percentages of total patients with completed tests are reported.

### Outcome Measures and Study Definitions

Respiratory co-infection was defined as collection of a respiratory culture, blood culture, or respiratory diagnostic (*Streptococcus pneumoniae* urine antigen, *Legionella* spp. urine antigen, or respiratory viral panel) positive for a respiratory pathogen within 72 hours of positive SARS-CoV-2 RT-PCR test collection. Respiratory co-infection was considered to have occurred if respiratory flora was grown from a respiratory culture and the patient was treated with systemic antibiotics. Other co-infection was defined as collection of a non-respiratory culture, positive for a non-respiratory pathogen within 72 hours of positive SARS-CoV-2 RT-PCR test collection. Subsequent infections were defined as those with collection of culture or diagnostic more than 72 hours after a positive SARS-CoV-2 RT-PCR test collection.

### Microbiologic and Laboratory Diagnostics

Patients with COVID-19 were identified via RT-PCR tests with Federal Drug Administration (FDA) Emergency Use Authorization (EUA) for detecting SARS-CoV-2 nucleic acid. RT-PCR samples were collected from nasopharyngeal and lower respiratory specimens. All diagnostic tests were performed according to the manufacturer’s package inserts. Influenza screening testing was performed using the BD Veritor System (BD Diagnostics, Sparks, MD). Molecular tests using the Verigene Respiratory Pathogens, Gram-Positive, and Gram-Negative Blood Culture nucleic acid tests (Nanosphere, Northbrook, IL, USA) were performed on nasopharyngeal samples and positive blood cultures, respectively. Nasopharyngeal screening for MRSA was done for patients receiving anti-MRSA antibiotic treatment utilizing Spectra MRSA (Thermo Fisher Scientific, Lenexa, KS, USA). Urine antigen testing for *S. pneumoniae* and *Legionella* spp. were performed using BinaxNow (Alere, Scarborough, ME). All testing for CDI was performed using the Illumigene molecular assay (Meridian Bioscience, Inc., Taunton, MA). All bacterial pathogens were identified via MicroScan WalkAway (Beckman Coulter, Inc., Brea, CA).

### Statistical Analysis

Descriptive analyses were performed for all variables. Mean ± standard deviation were determined for normally distributed variables and median and interquartile range were determined for non-normally distributed continuous variables. Count and proportions are presented for all categorical variables.

Bivariate comparisons using Chi-squared (or Fisher’s exact) tests were conducted for nominal data and two sample t-test or Mann-Whitney U test for continuous data (depending on normality distribution) were used to compare characteristics and outcomes between the sample of patients.

A Cox proportional hazards model was fit for time to death, controlling for treatment group and potential confounders (age in years, intensive care unit admission, mechanical ventilation, chronic obstructive pulmonary disease, coronary artery disease, chronic kidney disease, cirrhosis) based on a priori plausibility, bivariate associations within our data, and ruling out multicollinearity using variance inflation factors with values of less than 3 considered acceptable.

Significance was evaluated at α = .05 and all testing was 2-sided. Because of the potential for type I error due to multiple comparisons, findings for analyses of secondary end points should be interpreted as exploratory. All statistical analyses were performed using SPSS software (IBM SPSS Statistics, version 22.0; Chicago, IL, USA).

## Results

A total of 417 patients were screened with 289 patients ultimately included; 128 patients were excluded due to having an admission stay of less than 24 hours, with most being discharged from the emergency department. Among included patients, 48 (16.6%) had any co-infection (25 bacterial respiratory infections, 8.7%) compared to 241 (83.4%) without co-infection (Figure 1).

**Figure 1.**
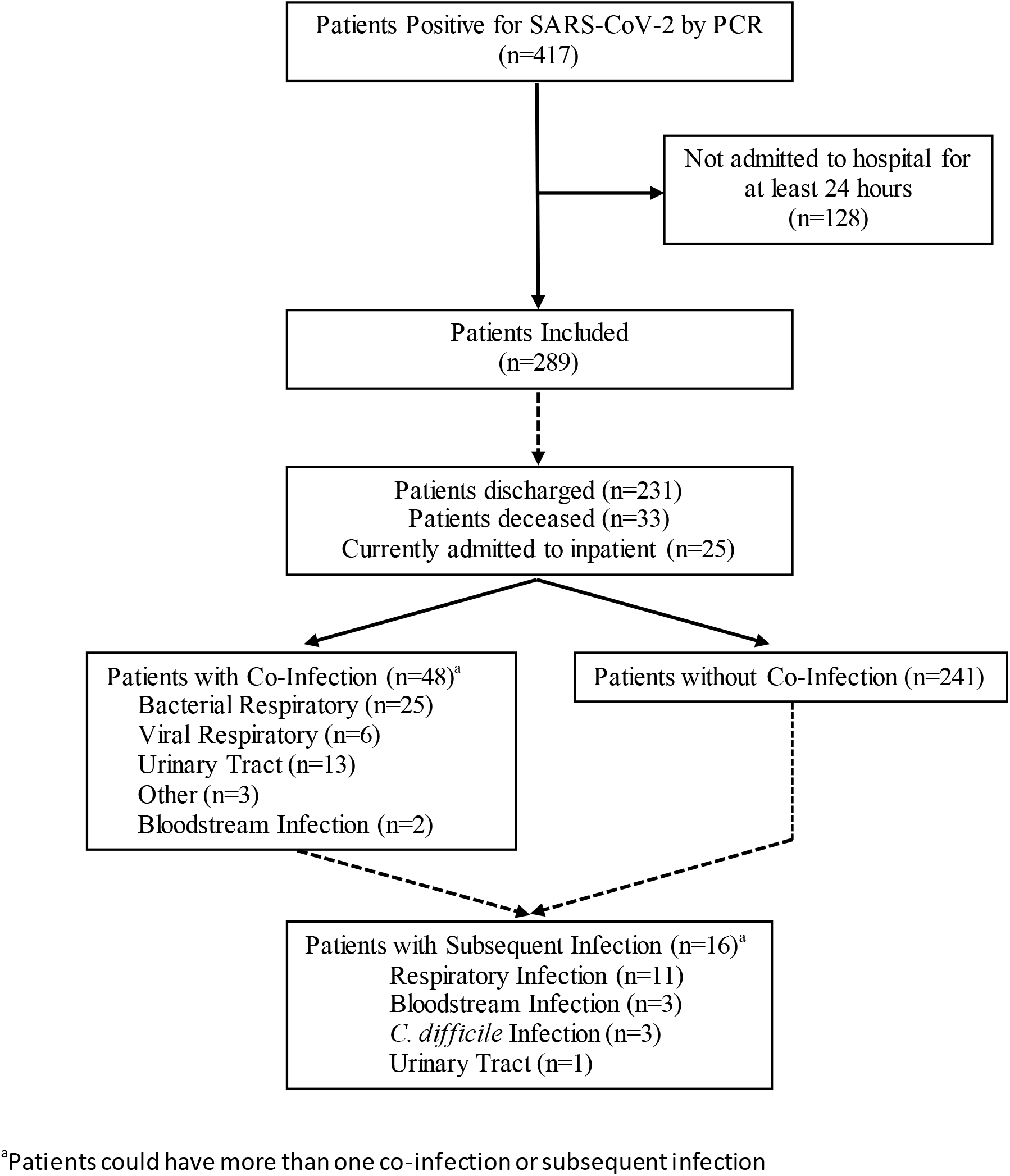
Flow Diagram of Patient Identification and Classification

Patient characteristics are described in Table 1. Mean age of included patients was 58.6 (SD +/−14.4) years and most had chest imaging consistent with pneumonia (86.1%). Additionally, most patients had at least one underlying comorbidity (81.7%) with diabetes (46.4%) and hypertension (61.6%) being the most common. No significant differences in comorbidities were observed between patients with a bacterial respiratory co-infection (BRC) and those without. Few patients were immunocompromised overall (10.3%) with no BRC patients found to have a history of transplant, active malignancy, or HIV. Among admission laboratory values assessed, median white blood cell count (WBC) was higher among patients with a BRC compared to those without (10.1 vs 7.0 ×10^9^/L, P = 0.002). Additionally, several other median laboratory values were higher among patients with a BRC compared to those without: lactate dehydrogenase (1210 vs 834, P = 0.008), C-reactive protein (182 vs 74, P = 0.013), procalcitonin (0.37 vs 0.07, P = 0.001), and interleukin-6 (48 vs 10, P = 0.011). Procalcitonin (PCT) greater than or equal to 0.25 had a sensitivity of 73.9% and a specificity of 65.2% for BRC. Whereas, at a PCT value greater than or equal to 0.5, sensitivity and a specificity were 43.5% and 81.3%, respectively.

**Table 1.** Characteristics, Laboratory Findings, Treatments, and Clinical Outcomes of 289 Hospitalized Patients with COVID-19

|  | **Total**<br>**(n=289)** | **Bacterial Respiratory**<br>**Co-Infection (n=25)** | **No Respiratory Co-**<br>**Infection (n=264)** | **P**<br>**value** |
| --- | --- | --- | --- | --- |
| Age, years | 58.6 +/-<br>14.4 | 59.1 +/- 15.9 | 58.5 +/- 14.3 |  |
| **Race/Ethnicity** |  |  |  |  |
| African American | 139 (48.3) | 7 (28) | 132 (50) |  |
| Hispanic | 68 (23.5) | 8 (32) | 60 (22.7) |  |
| Caucasian | 61 (21.1) | 7 (28) | 54 (20.5) |  |
| Other | 21 (7.3) | 3 (12) | 18 (6.8) |  |
| **Imaging** |  |  |  |  |
| Imaging showing<br>pneumonia, n (%) | 242/281<br>(86.1) | 23 (92.0) | 219 (83.0) |  |
| Chest radiograph | 218/275<br>(79.3) | 22 (88.0) | 196 (74.2) |  |
| CT | 115/131<br>(87.8) | 16 (64.0) | 99 (37.5) |  |
| **Comorbidities** |  |  |  |  |
| Asthma | 26 (9.0) | 3 (12.0) | 23 (8.7) |  |
| Chronic Heart Failure | 19 (6.6) | 1 (4.0) | 18 (6.8) |  |
| Chronic Obstructive<br>Pulmonary Disease | 18 (6.2) | 2 (8.0) | 16 (6.1) |  |
| Chronic Kidney Disease | 21 (7.3) | 0 (0) | 21 (8.0) |  |
| Cirrhosis | 3 (1.0) | 0 (0) | 3 (1.1) |  |
| Coronary artery disease | 35 (12.1) | 3 (12.0) | 32 (12.1) |  |
| Diabetes | 134 (46.4) | 13 (52.0) | 121 (45.9) |  |
| Hypertension | 178 (61.6) | 16 (64.0) | 162 (61.4) |  |
| End-stage renal disease | 18 (6.2) | 2 (8.0) | 16 (6.1) |  |
| Human Immunodeficiency<br>Virus | 7 (2.4) | 0 (0) | 7 (2.7) |  |
| Malignancy | 20 (6.9) | 0 (0) | 20 (7.6) |  |
| Sleep apnea | 5 (1.7) | 1 (4.0) | 4 (1.5) |  |
| Transplant | 3 (1.0) | 0 (0) | 3 (1.1) |  |
| No PMH | 53 (18.3) | 3 (12.0) | 50 (18.9) |  |
| **Laboratory Values on Admission** |  |  |  |  |
| WBC (x10<sup>9</sup>/L) | 7.1 (5.1,<br>9.6) | 10.1 (6.5, 12.5) | 7.0 (4.9, 9.3) | 0.002 |
| Absolute Lymphocytes<br>(x10<sup>9</sup>/L) | 1.0 (0.7,<br>1.4) | 1.1 (0.5, 1.4) | 1.0 (0.7, 1.4) | 0.617 |
| Lactate Dehydrogenase<br>(U/L) | 846 (603,<br>1210) | 1210 (755, 1451) | 834 (589, 1167) | 0.008 |
| Ferritin (ng/mL) | 411 (192,<br>908) | 510 (368, 901) | 372 (182, 916) | 0.267 |
| C-Reactive Protein<br>(mg/dL) | 76 (39,<br>189) | 182 (58, 266) | 74 (38, 181) | 0.013 |
| Procalcitonin (ng/mL) | 0.16 (0.08,<br>0.42) | 0.37 (0.19, 1.53) | 0.07 (0.15, 0.38) | 0.001 |
| Procalcitonin >0.25 | 95 (32.9) | 17 (68.0) | 78 (29.5) | <.001 |
| Procalcitonin >0.5 | 54 (18.7) | 10 (40.0) | 44 (16.7) | 0.004 |
| Interleukin-6 (pg/mL) | 11 (<5, 37) | 48 (10, 71) | 10 (5, 21) | 0.011 |
| D-Dimer (ng/mL) | 1.16 (0.71, 2.35) | 1.51 (0.96, 3.87) | 1.13 (0.66, 2.25) | 0.114 |
| **Treatments** |  |  |  |  |
| Antibiotic | 271 (93.8) | 25 (100) | 246 (93.2) | 0.381 |
| Hydroxychloroquine | 163 (56.4) | 18 (72) | 145 (54.9) | 0.151 |
| Remdesivir | 28 (9.7) | 0 (0) | 28 (10.6) | <0.001 |
| Steroids | 70 (24.2) | 12 (48.0) | 58 (22.0) | 0.004 |
| Tocilizumab | 18 (6.2) | 5 (20.0) | 13 (4.9) | 0.013 |
| **Clinical Outcomes\*** |  |  |  |  |
| Length-of-stay | 7 (4, 11) | 10 (5, 16) | 7 (4, 11) | 0.124 |
| ICU admission | 105 (36.3) | 21 (84.0) | 84 (31.8) | <.001 |
| ICU Length-of-stay | 7.5 (4, 16.5) | 8.5 (5, 12) | 7 (3, 17) | 0.805 |
| Mechanical ventilation | 81 (28.0) | 18 (72.0) | 63 (23.9) | <.001 |
| Mechanical ventilation duration, days | 9 (5, 14) | 6.5 (5, 12) | 9.5 (5, 15) | 0.253 |
| In-hospital mortality | 33 (11.4) | 9 (45.0) | 24 (9.8) | <.001 |

Age is presented as mean +/- SD. Other continuous variables are presented as median (IQR) and compared with Mann-Whitney U test. Categorical variables are shown as number (%). Chi-squared or Fisher's exact tests were used to compare categorical variables between groups.
\*Information not available or incomplete for patients remaining admitted

Some therapies targeting COVID-19 were more commonly administered to patients with a BRC compared to those without including systemic corticosteroids (48 vs 22%, P = 0.004), and tocilizumab (20 vs 4.9%, P = 0.013). In contrast, patients without a BRC received remdesivir more often (10.6% vs 0, P =<0.001). There were no observed differences in rates of administration of hydroxychloroquine. Antibiotics were given to most patients (93.8%) with no differences between groups observed.

Blood cultures were obtained from most patients overall (242/289, 83.7%) and were commonly obtained among patients with a BRC compared to those without (100 vs 82.2%, P = 0.019) (Table 2). Other microbiologic diagnostics were also more frequently completed in patients with a BRC compared to those without including: influenza testing (96 vs 72%), *Streptococcus pneumoniae* urine antigen testing (80 vs 52.7%), MRSA nasopharyngeal screening (48 vs 22.3%), and respiratory cultures (64 vs 14.8%). Conversely, *Legionella* urine antigen and *C. difficile* testing were similar amongst groups.

**Table 2.** Microbiologic Diagnostic Evaluation, by Bacterial Respiratory Co-Infection Status

| **n (%)** | **Total (n=289)** | **Bacterial Respiratory Co-Infection (n=25)** | **No Bacterial Respiratory Co-Infection (n=264)** |
| --- | --- | --- | --- |
| **Blood Cultures** | 242 (83.7) | 25 (100) | 217 (82.2) |
| **Flu (Any)** | 214 (74.0) | 24 (96.0) | 190 (72.0) |
| **Flu Screen** | 182 (63.0) | 22 (88.0) | 160 (60.6) |
| **Flu PCR** | 198 (68.5) | 24 (96.0) | 174 (65.9) |
| **Strep pneumo Ur Ag** | 159 (55.0) | 20 (80.0) | 139 (52.7) |
| **Legionella Ur Ag** | 28 (9.7) | 3 (12.0) | 25 (9.5) |
| **RVP** | 197 (68.2) | 24 (96.0) | 173 (65.5) |
| **MRSA nasal screen** | 71 (24.6) | 12 (48.0) | 59 (22.3) |
| **Respiratory Culture (Any)** | 55 (19.0) | 16 (64.0) | 39 (14.8) |
| **Tracheal aspirate** | 42 (14.5) | 12 (48.0) | 30 (11.4) |
| **Bronchoscopy** | 4 (1.4) | 0 (0) | 4 (1.5) |
| **Sputum** | 16 (5.5) | 5 (20.0) | 11 (4.2) |
| **CDI Testing** | 32 (11.1) | 2 (8.0) | 30 (11.4) |

Among patients with a respiratory co-infection, 25 (91.4%) had a bacterial pathogen and 6 (8.6%) had a respiratory virus other than SARS-CoV-2 (Table 3). Among bacterial pathogens, respiratory flora was most commonly identified (n = 15, 60%) followed by *Staphylococcus aureus* (n = 5, 20%) and *S. pneumoniae* (n = 3, 12%). Of the *S. aureus* isolates, 3/5 (60%) were methicillin-resistant. No predominant respiratory virus was identified as causing co-infection. As co-infection rates were low, few positive microbiologic diagnostics were observed overall. Of note, only 3/159 (1.8%) of *S. pneumoniae* urine antigens, 0/ 28 *Legionella* urine antigens, 7/197 (3.6%) of respiratory viral panels, and 1/71 (1.4%) nasopharyngeal MRSA screens performed were positive.

**Table 3.** Pathogens Identified, by Infection Type

| Infection Type |  | Pathogen | N |
| --- | --- | --- | --- |
| **Respiratory Co-Infection (n=31)** |  |  |  |
|  | Bacterial (n=25) | *Bordetella holmesii* | 1 |
|  |  | *Staphylococcus aureus* | 5 |
|  |  | *Streptococcus pneumoniae* | 3 |
|  |  | Group A Streptococci | 1 |
|  |  | Oral Bacterial Flora | 15 |
|  | Virus (n=6) | Adenovirus | 1 |
|  |  | Influenza A | 1 |
|  |  | Influenza B | 1 |
|  |  | Metapneumovirus | 1 |
|  |  | Rhinovirus | 2 |
| **Subsequent Respiratory Infection (n=11)** |  |  |  |
|  | Bacterial (n=10) | *Staphylococcus aureus* | 2 |
|  |  | *Streptococcus pneumoniae* | 1 |
|  |  | *Pseudomonas aeruginosa* | 3 |
|  |  | *Klebsiella* spp. | 2 |
|  |  | *E. coli* | 1 |
|  |  | *Stenotrophomonas maltophilia* | 1 |
|  | Fungal (n=1) | *Aspergillus niger* | 1 |
| **Other Subsequent Infections (n=7)** |  |  |  |
|  | Bloodstream (n=3) | *Staphylococcus epidermidis* | 1 |
|  |  | *Candida* sp. | 2 |
|  | Urinary Tract (n=1) | Group B Streptococci | 1 |
|  | Other (n=3) | *C. difficile* | 3 |

\*Patient could have more than one pathogen or infection

Hospital outcomes are presented in Table 1, noting 25 (8.7%) patients were still hospitalized at the time of final analysis. ICU admission was common overall (36.3%) and occurred more in patients with a BRC compared to those without (84 vs 31.8%, P=< 0.001). Moreover, patients with a BRC were more likely to be mechanically ventilated (72 vs 23.9%, P=<0.001) compared to those without. In-hospital mortality was higher among patients with a BRC compared to those without in an unadjusted bivariate analysis (45 vs 9.8%; P=<0.001). Among subgroup of patients admitted to the ICU, in-hospital mortality remained higher in the group with a BRC (9/16, 56.3%) compared to those without (20/66, 30.3%; P=0.051). No differences in ICU admission, mechanical ventilation, or in-hospital mortality was observed for patients with a non-respiratory co-infection or those with a viral respiratory co-infection compared to the non-BRC group as a whole.

In a Cox proportional hazards regression, following adjustment for age, ICU admission, mechanical ventilation, corticosteroid administration, and pre-existing comorbidities, patients with a BRC had an increased risk for in-hospital mortality (adjusted HR, 3.37; 95% CI, 1.39 to 8.16; P = 0.007) (Figure 2, Table 4).

**Figure 2.**
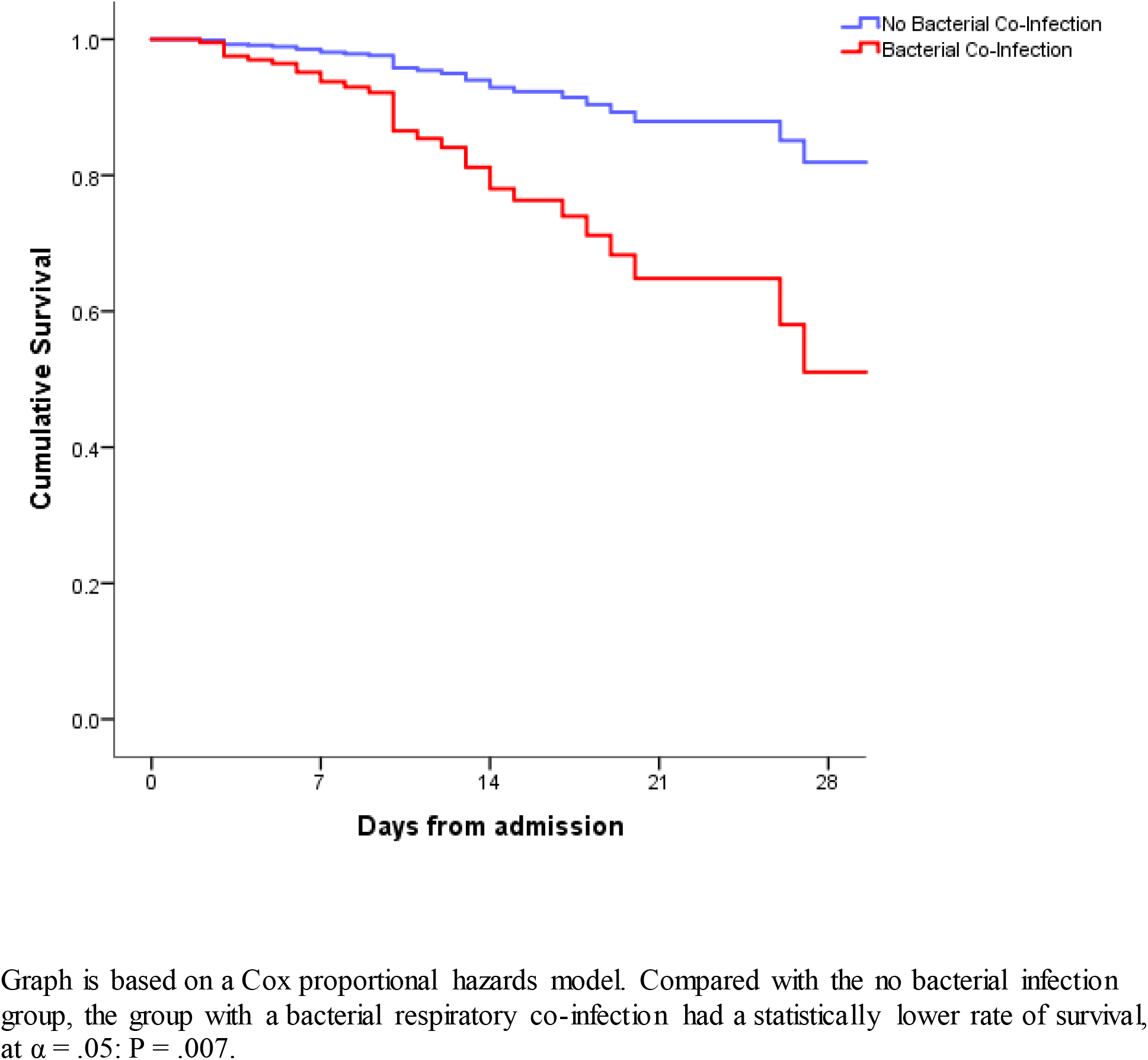
Model-Adjusted Estimated In-Hospital Mortality, by Bacterial Respiratory Co-Infection Status

**Table 4.** Variables retained in Cox Proportional Hazards Model for In-Patient Mortality

| **Covariate** | **aHR (95% CI)** | ***P* value** |
| --- | --- | --- |
| **Age, years** | 1.044 (1.011-1.079) | 0.009 |
| **ICU admission** | 6.341 (1.498-26.851) | 0.012 |
| **Mechanical ventilation** | 1.146 (0.373-3.524) | 0.812 |
| **COPD** | 1.218 (0.473-3.137) | 0.683 |
| **CAD** | 1.151 (0.416-3.184) | 0.786 |
| **CKD** | 1.415 (0.469-4.266) | 0.538 |
| **Cirrhosis** | 5.912 (1.104-31.669) | 0.038 |
| **Bacterial respiratory co-infection** | 3.368 (1.391-8.155) | 0.007 |
| **Steroids** | 0.333 (0.142-0.783) | 0.012 |

Abbreviations: CAD, coronary artery disease; CKD, Chronic kidney diseases; COPD, Chronic obstructive pulmonary disease;

A total of 21 subsequent infections occurred in 16 (5.5%) of patients; respiratory infections were most common (n = 12, 57.1%) followed by bloodstream infections (n = 3, 14.3%) and *C. difficile* infections (n = 3, 14.3%) (Table 3). Time from admission to subsequent infection ranged from 5 to 23 days (median of 11 days). Patients that received systemic corticosteroids were more likely to develop a subsequent infection (11/70, 15.7%) during hospitalization than those that did not (5/219, 2.3%; P = <0.001). Moreover, patients that received systemic corticosteroids were more likely to develop a subsequent respiratory infection (7/70, 10%) compared to those that did not (4/219, 1.8%; P = 0.002). Each of the 3 patients with subsequent fungal infections had received systemic corticosteroids. Similar to corticosteroids, patients that received tocilizumab were more likely to have a subsequent infection (5/18, 27.8%) than those that did not (11/271, 4.1%; P = 0.001).

## Conclusions

Among hospitalized patients with COVID-19, co-infections occurred in only 16.6% of patients. Moreover, respiratory co-infections with bacterial (8.7%) or viral (2.1%) pathogens were infrequent and no predominant pathogens were identified. For non-BRC (sources other than respiratory) or viral co-infections, there was no increased morbidity or mortality observed. When BRC did occur they were associated with higher rates of ICU admission, mechanical ventilation, and in-hospital mortality. Following adjustment for age, ICU admission, mechanical ventilation, corticosteroid administration, and pre-existing comorbidities, patients with a BRC had an increased risk for in-hospital mortality (adjusted HR, 3.37; 95% CI, 1.39 to 8.16; P = 0.007). Subsequent infections occurred in 5.5% (n = 16) patients, with onset ranging from 5 to 23 days after admission (median 11 days). Subsequent infections were most commonly of respiratory source (57.1%) followed by bloodstream and CDI (14.3% each). Patients that received either corticosteroids or tocilizumab were more likely to have developed a subsequent infection.

Unlike previous observations with other respiratory viruses causing pneumonia, SARSCoV-2 does not appear to have high rates of co-infection with either bacterial or other viral pathogens.[10] This contrast may be attributable to the widespread prevalence of the disease and presentation to the hospital with the disease in patients normally considered immunocompetent compared to other respiratory viruses. Several case reports have demonstrated that co-infections with respiratory viruses, including influenza, and SARS-CoV-2 do occur.[7, 11–21] Recently an evaluation of 5,700 hospitalized patients with COVID-19 in the New York City area similarly showed only 42 of 1996 patients tested (2.1%) with another respiratory virus identified on admission.[8] One analysis of patients presenting to an emergency department showed that 24 of 116 (20.7%) patients with COVID-19 had a viral co-infection detected, however, only 1 of the 24 was admitted to the hospital.[15] It is possible that there is some disparity between rates of viral co-infection among patients seen in the inpatient and outpatient settings as such patients are likely to vary in age and comorbidities. Moreover, respiratory viruses are notably seasonal and the timing of our evaluation (influenza activity was declining by the time of first SARS-CoV-2 detection) and others is therefore notable as it relates to viral co-infections with the potential for variance to occur depending on circulating respiratory viruses. Bacterial co-infection was also infrequently identified with SARS-CoV-2 in this study cohort.

The low rates of bacterial co-infections identified are noteworthy as they contrast sharply with antibiotic exposures observed. Rawson and colleagues conducted a review of the early data related to bacterial and fungal co-infections with SARS-CoV-2 and found that 72% of patients with COVID-19 received systemic antibacterials yet only 62 of 806 (8%) had an identified bacterial or fungal co-infection. We similarly found high rates of antibacterial administration among hospitalized patients with COVID-19 (93.8% of patients) and low rates of bacterial co-infection (14.5% with any bacterial co-infection and 8.6% with respiratory co-infection). Although BRC rates with SARS-CoV-2 are seemingly lower than those reported with influenza (ranging from 11 to 35% in most reports),[22] increased morbidity and mortality is still of concern. Previously, associations between BRC related to 2009 pandemic influenza and higher rates of mechanical ventilation and mortality have been described.[5] We similarly observed higher rates of mechanical ventilation and in-hospital mortality among patients with BRC. Synergistic interactions between bacterial pathogens such as *S. pneumoniae* have been described and the pathophysiologic mechanisms could also hold true for SARS-CoV-2.[6, 23, 24]

To our knowledge, there is currently no specific antimicrobial stewardship interventions related to COVID-19 reported. A meta-analysis evaluating the ability for procalcitonin to distinguish viral from bacterial pneumonia demonstrated sensitivity and specificity of serum procalcitonin were 0.55 and 0.76, respectively.[25] This performance likely varies to some degree on patient setting,[26, 27] but is fairly consistent with our findings for procalcitonin and BRC (sensitivity of 73.9% and specificity of 65.2% at a cut-off value of 0.25 ng/mL). Utilization of procalcitonin could prove helpful in decreasing unnecessary and potentially harmful antibiotic utilization in treating hospitalized COVID-19 patients and warrants further investigation.

Diagnostic tests needed to effectively evaluate hospitalized COVID-19 patients for co-infection could also play a substantial role in optimizing management. Schimmel et al. reported on utilization of *S. pneumoniae* urinary antigen utilization among 159, 894 hospital admissions for community-acquired pneumonia in the U.S. and determined that less than 20% of admissions had the test performed.[28] Moreover, there appeared to be opportunity for positive tests to lead to reductions in broad-spectrum antibiotic use. More than half (55%) of patients in the current study had a *S. pneumoniae* urine antigen performed, but only 3 resulted positive. This test could have helpful implications related to personal protective equipment (PPE) and exposure minimization, but the low yield of the test should be balanced with its potential impact.

Several limitations of this study should be recognized. First, the evaluation was conducted at 4 hospitals within the same healthcare system in Dallas, Texas, USA during the first two months in which the SARS-CoV-2 pandemic was recognized in the area and application to other geographic areas or time periods may not apply. Specifically, the seasonality of other respiratory viruses may alter rates of co-infection observed and continued observations over time, notably in Fall and Winter months, may help in delineating seasonal trends. Second, the decision to consider patients with only respiratory flora as having a BRC could be considered a limitation in interpreting the association of BRC on clinical outcomes among patients with COVID-19. However, when analyses excluding these patients were done, rates of ICU admission, mechanical ventilation, and in-hospital mortality remained higher among patients with a BRC suggesting that these patients are similar to those with more obvious BRC (e.g., *S. pneumoniae*). Moreover, respiratory flora and disruption of the lung microbiome are increasingly recognized as potential causes of pneumonia and could explain the “missing” pathogens frequently observed.[29–32] Molecular diagnostic tests for respiratory specimen could also prove fruitful in increasing yield of pneumonia pathogens, but were not available during this study. Third, PPE restrictions to minimize healthcare exposure may impact obtainment and/or quality or frequency of respiratory cultures or other diagnostics; thus potentially influencing the rates of co-infection identified. Fourth, it is difficult to separate the impact of BRC and other factors such as COVID-19 severity or underlying disease states on clinical outcomes. Therefore, the findings of worse clinical outcomes among patients with BRC and COVID-19 should be strictly seen as hypothesis generating. Additionally, we did not address the potential efficacy of antibiotic therapy herein, leaving it unknown what the impact of antibiotic therapy on patients with BRC would be. Finally, 25 patients remained hospitalized at the time of follow-up completion and it is unknown how rates or types of subsequent infections in this study might be affected if longer follow-up was possible among this group.

Co-infections with SARS-CoV-2 are uncommon among hospitalized patients; however, BRC is associated with worse clinical outcomes including higher mortality when it does occur. Future evaluations on antibiotic therapy and antimicrobial stewardship are warranted to determine how to minimize use of unnecessary antimicrobials while still optimizing therapy among patients with co-infections. Additionally, evaluations of necessary diagnostic work-up among hospitalized COVID-19 patients could assist clinicians in decision-making and minimize exposure to this highly contagious virus. SARS-CoV-2 and COVID-19 are likely to remain a primary cause of hospitalization in the coming months to years and an increased understanding of the disease-state continuum and its management is needed to improve clinical outcomes associated with this severe illness.

## Data Availability

All data collected for the referenced study is maintained and available for the next 7 years by the authors.

## Notes

**Competing Interests:** All authors have completed the ICMJE uniform disclosure form at http://www.icmje.org/coi_disclosure.pdf and declare: no support from any organization for the submitted work; MPC has received honorariums from Nabriva Therapeutics, Theravance Biopharma, Inc., Paratek Pharmaceuticals, and bioMerieux, Inc., outside the submitted work; EAD reports being a sub-investigator for clinical trials through Gilead, research funding from Nektar, personal fees for delivering educational presentations for Sanofi-Pasteur, and has received honorariums from Cumberland Pharmaceuticals and Tetraphase Pharmaceuticals, outside the submitted work; no other authors report relationships or activities that could appear to have influenced the submitted work.

### Competing Interest Statement

All authors have completed the ICMJE uniform disclosure form at www.icmje.org/coi_disclosure.pdf and declare: no support from any organization for the submitted work; MPC has received honorariums from Nabriva Therapeutics, Theravance Biopharma, Inc., Paratek Pharmaceuticals, and bioMerieux, Inc., outside the submitted work; EAD reports being a sub-investigator for clinical trials through Gilead, research funding from Nektar, personal fees for delivering educational presentations for Sanofi-Pasteur, and has received honorariums from Cumberland Pharmaceuticals and Tetraphase Pharmaceuticals, outside the submitted work; no other authors report relationships or activities that could appear to have influenced the submitted work.

### Funding Statement

No support from any organization for the submitted work.

### Author Declarations

The Aspire institutional review board approved this multicenter observational cohort study as minimal-risk research using data collected for routine clinical practice and waived the requirement for informed consent.

